# Direct Comparison of Antibody Responses to Four SARS-CoV-2 Vaccines in Mongolia

**DOI:** 10.1101/2021.08.22.21262161

**Authors:** Naranjargal J. Dashdorj, Oliver F. Wirz, Katharina Röltgen, Emily Haraguchi, Anthony S. Buzzanco, Mamdouh Sibai, Hannah Wang, Jacob A. Miller, Daniel Solis, Malaya K. Sahoo, Sumiya Byambabaatar, Purevjargal Bat-Ulzii, Anir Enkhbat, Enkhtuul Batbold, Delgersaikhan Zulkhuu, Byambasuren Ochirsum, Tungalag Khurelsukh, Ganbold Dalantai, Natsagdorj Burged, Uurtsaikh Baatarsuren, Nomin Ariungerel, Odgerel Oidovsambuu, Andreas S. Bungert, Zulkhuu Genden, Dahgwahdorj Yagaanbuyant, Altankhuu Mordorj, Theodore Jardetzky, James L. Wilbur, Jacob N. Wohlstadter, George B. Sigal, Benjamin A. Pinsky, Scott D. Boyd, Naranbaatar D. Dashdorj

## Abstract

Different vaccines for SARS-CoV-2 are approved in various countries, but few direct comparisons of the antibody responses they stimulate have been reported. We collected plasma specimens in July 2021 from 196 Mongolian participants fully vaccinated with one of four Covid vaccines: Pfizer/BioNTech, AstraZeneca, Sputnik V and Sinopharm. Functional antibody testing with a panel of nine SARS-CoV-2 viral variant RBD proteins reveal marked differences in the vaccine responses, with low antibody levels and RBD-ACE2 blocking activity stimulated by the Sinopharm and Sputnik V vaccines in comparison to the AstraZeneca or Pfizer/BioNTech vaccines. The Alpha variant caused 97% of infections in Mongolia in June and early July 2021. Individuals who recover from SARS-CoV-2 infection after vaccination achieve high antibody titers in most cases. These data suggest that public health interventions such as vaccine boosting, potentially with more potent vaccine types, may be needed to control the COVID-19 pandemic in Mongolia and worldwide.

## Main Text

Several different vaccines for SARS-CoV-2 have been approved for use in various countries and are being actively deployed in an effort to combat the COVID-19 pandemic, but few direct comparisons of the antibody responses they stimulate have been reported. Many viral variants have arisen since the initial months of the pandemic, and are in circulation with different geographical distributions and susceptibility to antibody responses elicited to Wuhan-Hu1 antigens (Garcia-Beltran et al., 2021; Hoffmann et al., 2021; Muik et al., 2021; Planas et al., 2021a; Röltgen et al., 2021; Supasa et al., 2021). Breakthrough infections with SARS-CoV-2 in previously vaccinated individuals, together with data from the clinical trials supporting regulatory approval of the vaccines indicate that there are disparities in the amount of protection against infection that they provide (Khoury et al., 2021; Wall et al., 2021). Beginning in February 23, 2021, Mongolia carried out a vigorous campaign of vaccination of its citizens, and achieved a high rate of 61.4% of the total population fully vaccinated, with an additional 6.3% having received a single dose. The adult population has primarily been vaccinated with the Sinopharm vaccine (89.2% of vaccinated adults). In recent months, widespread outbreaks of SARS-CoV-2 infection have been reported in Mongolia, with 86 cases per 100,000 in mid-June 2021 at the height of the outbreak, decreasing to approximately 40 cases per 100,000 at the end of July 2021, including many vaccinated individuals. The viral variants responsible for these infections are currently unknown.

We collected plasma specimens in a five-day period from July 3 to 7, 2021 from Mongolian participants who had been fully vaccinated with one of four Covid vaccines: Pfizer/BioNTech (BNT162b2), AstraZeneca (ChAdOx1-S), Sputnik V (Gam-COVID-Vac) and Sinopharm (BBIBP-CorV). Participants were recruited by public announcement and volunteers were enrolled after signing the consent form approved by the Ethics Review Board at the Ministry of Health of Mongolia. (**Supplemental Material**). Antibodies were analyzed in the plasmas of 196 participants divided between the vaccine groups (47, 50, 45 and 54 individuals for Pfizer/BioNTech, AstraZeneca, Sputnik V and Sinopharm, respectively) and selected to balance age, sex and time after second vaccine dose (**Figs. S1, S2**). We measured antibody blocking of angiotensin-converting enzyme 2 (ACE2) host receptor protein binding to SARS-CoV-2 spike receptor binding domains (RBD) from nine viral variants of concern or interest according to CDC and WHO definitions, using an electrochemiluminescence assay platform from Meso Scale Discovery. The RBDs that were tested were (with RBD amino acid changes from Wuhan-Hu1 in parentheses): Alpha (N501Y), Beta (K417N, E484K, N501Y), Gamma (K417T, E484K, N501Y), Delta (L452R, T478K), Epsilon (L452R), Eta/Iota/Zeta (E484K), Kappa (L452R, E484Q), B.1.526.2 (S477N) and P.3 (E484K, N501Y), as well as Wuhan-Hu1. Antibody blocking of ACE2 binding to each RBD for each vaccine type is shown in **Fig. 1A**. RBD-ACE2 blocking antibody results for participants of different ages (<60 or ≥60 years) and sex are shown in **Fig. 1B**. We additionally measured the titers of IgG antibodies binding to RBD, spike (S) and nucleocapsid (N) antigens of Wuhan-Hu1 SARS-CoV-2 in all specimens (**Fig. S3A**). Results from ACE2 blocking antibody assays and anti-RBD IgG binding assays have been shown to be highly correlated with neutralizing antibody titers from pseudotyped viruses displaying SARS-CoV-2 spike, and authentic SARS-CoV-2 virus (Feng et al., 2021; Röltgen et al., 2020, 2021). We reconfirmed these previously reported correlations between RBD-ACE2 blocking antibody results, anti-RBD IgG titers and, for a subset of samples, a pseudotyped lentivirus neutralization assay with Wuhan-Hu1 spike protein (**Fig. S3B**).

**Figure 1.**
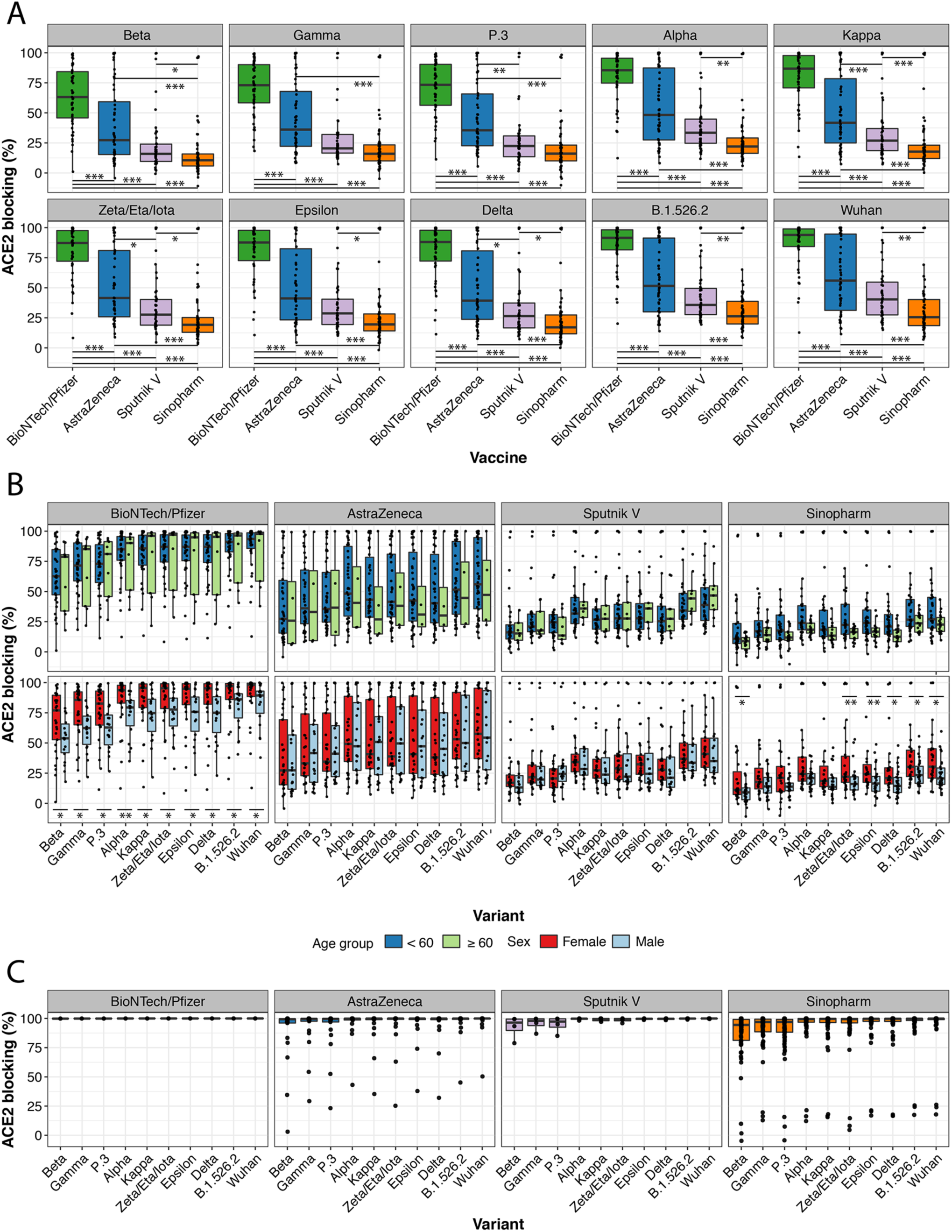
Vaccine-induced antibody blocking of RBD-ACE2 binding for different viral variants. (A) Percentage blocking of ACE2 binding to RBD of specified viral variants by plasma antibodies of recipients of Pfizer/BioNTech, AstraZeneca, Sputnik V and Sinopharm vaccines is shown. Significance of differences between groups was calculated by pairwise Wilcoxon test with Bonferroni correction (*, **, *** indicate P<0.05, P<0.01, P<0.001 respectively). (B) Blocking antibody responses stratified by participant age (<60 years, or ≥ 60 years) and sex. (C) RBD-ACE2 blocking antibody responses for 99 participants with confirmed SARS-CoV-2 infection post-vaccination with the indicated vaccines.

ACE2 blocking antibody activity was strikingly different between the vaccines tested. Within each vaccine group, differences were also observed in antibody activity for the different viral variant antigens, although these were smaller than the differences between the vaccine groups. The Pfizer/BioNTech vaccine elicited the strongest ACE2 blocking antibody activity, followed by AstraZeneca vaccine, then Sputnik V, with the lowest levels from Sinopharm (**Fig. 1A**). Differences between the vaccine responses were highly significant for most viral variants, although differences between the Sputnik V and AstraZeneca did not always reach significance. ACE2 blocking antibody activity for viral variants of concern or interest showed a consistent hierarchy of decreased blocking, with the greatest decrease for the Beta, Gamma and P.3 variants and more modest decreases for the other variants (**Fig. 1A**), similar to previously reported results (Garcia-Beltran et al., 2021; Röltgen et al., 2021). There was greater variation between participants in the ACE2 blocking antibody responses within the AstraZeneca group compared to the other vaccines. Median times of sampling post-second vaccination were between two and three months for all vaccines (**Fig. S1**); we note some Sputnik V recipient plasmas were collected at later time points after the second vaccine dose compared to the other vaccines, but plotting ACE2 blocking indicated that this had little effect on the results (**Fig. S2**). Titers of IgG specific for RBD and spike showed similar results to the ACE2 blocking antibody results, with decreasing titers from Pfizer/BioNTech to AstraZeneca to Sputnik V to Sinopharm (**Fig. S3A**). The age of vaccine recipients and proportions of males and females in each group were comparable (**Fig. S1)**. Notably, males had significantly lower ACE2 blocking antibody values than females to the Pfizer/BioNTech and Sinopharm vaccines, but not to the AstraZeneca and Sputnik V vaccines (**Fig. 1B**).

Testing for antibodies to SARS-CoV-2 nucleocapsid (N) antigen assessed for evidence of prior infection, since the Pfizer/BioNTech mRNA vaccine, and the AstraZeneca and Sputnik V adenoviral vectored vaccines do not contain or produce N antigen. Most recipients of the Sinopharm vaccine, which contains inactivated SARS-CoV-2 viruses, showed the expected increased levels of anti-N antibodies compared to other vaccine recipients (**Fig. S3A**), although most were below the cutoff for seroconversion in this assay. The anti-N IgG assay results also identified some participants with evidence of prior infection amongst the other vaccine recipients who had no reported history of infection (5, 8 and 4 individuals for Pfizer/BioNTech, AstraZeneca and Sputnik V, respectively). Very high amounts of anti-N IgG well above the cut-off for seroconversion were observed in three Sinopharm individuals. Pre-vaccination specimens were not available for participants to further evaluate evidence of infection prior to vaccination, but the Sinopharm and Sputnik V recipients with the highest anti-N antibodies had significantly higher ACE2-blocking antibody activity than others in their vaccination groups, suggesting that these individuals had a combination of infection and vaccination. To further evaluate the serological effects of combined SARS-CoV-2 infection and vaccination, an additional cohort of 99 participants who had been vaccinated and then had documented SARS-CoV-2 infections (1, 21, 4 and 73 recipients of Pfizer/BioNTech, AstraZeneca, Sputnik V and Sinopharm vaccines, respectively) was analyzed with the RBD-ACE2 blocking antibody assay (**Fig. 1C**). ACE2 blocking antibody activity was very high in almost all of these individuals, with a median of >99% blocking against all variants except the Beta, Gamma and P.3 variants. Small numbers of AstraZeneca and Sinopharm recipients showed lower blocking activity against most variants after testing positive for infection post-vaccination (**Fig. 1C**).

It is important to note that ACE2 blocking antibody assay results are only a surrogate for potential protection of the individual from infection by SARS-CoV-2, as are the results from other assays such as viral neutralization assays. At present, there is no generally accepted threshold for results in any of these assays that can serve as a correlate of protection from infection by SARS-CoV-2. Initial attempts to evaluate decreases in risk of SARS-CoV-2 infection following vaccination suggest that the results of surrogate assays such as anti-RBD or anti-spike IgG measurement, as well as neutralization assays are correlated with protection against symptomatic infection (Feng et al., 2021).

The serological data from recipients of the four vaccines tested suggested that Sinopharm recipients, who comprise 89.2% of vaccinated adults in Mongolia, as well as the smaller number of individuals vaccinated with Sputnik V or AstraZeneca vaccines, could be particularly susceptible to breakthrough infections. To assess whether viral variants that are more resistant to antibodies elicited by Wuhan-Hu1 antigens are responsible for the ongoing wave of breakthrough infections in Mongolia, we carried out viral genotyping with spike N501Y, E484K, and L452R mutation-specific RT-qPCR on 182 nasopharyngeal swabs collected between June 18 and July 5, 2021 from individuals with Sinopharm post-vaccination SARS-CoV-2 infection. Viral genotyping showed that these infections were dominated by presumptive Alpha variants with the N501Y mutation, accounting for 97.3% (177/182) of cases tested. The other samples were comprised of two samples with the L452R mutation, consistent with several lineages including the Delta variant, and three samples without N501Y, E484K, or L452R mutations, unlikely to represent variants of concern or interest. Samples containing N501Y were further tested for spike del69_70 by RT-qPCR; this characteristic deletion was detected in all (177/177) of these samples, confirming that Sinopharm post-vaccination cases were caused primarily by the Alpha variant. The Alpha variant has minimal evasion of antibody responses elicited by Wuhan-Hu1 antigens (Muik et al., 2021) (**Fig. 1A**), suggesting that the breakthrough infections in Mongolia are related to the overall low antibody levels, rather than being caused by viral variants with greater antibody evasion capabilities.

In summary, this direct comparison of vaccine-elicited functional antibody responses to a panel of nine SARS-CoV-2 viral variant RBD proteins indicates that there are marked differences in the serological responses generated by each vaccine, with relatively low antibody titers and ACE2 blocking activity stimulated by the Sinopharm and Sputnik V vaccines, variable levels for the AstraZeneca vaccine, and the highest values for the Pfizer/BioNTech vaccine. Most individuals who recover from infection with SARS-CoV-2 after receiving any of the vaccines studied show elevated ACE2 blocking antibody activity, comparable to that seen in uninfected recipients of the Pfizer vaccine. Breakthrough infections in Mongolia in June and July 2021 are largely attributable to the more infectious, but not highly immune-evasive Alpha variant (Davies et al., 2021; Planas et al., 2021b). Limitations of this study include its retrospective observational design without randomized assignment of individuals to vaccine groups, the lack of pre-vaccination plasma samples, and the lack of plasma specimens following vaccination but before infection in the 99 individuals with documented breakthrough infections. The data indicate that there are major differences in the magnitude of functional antibody responses stimulated by the four vaccines studied, and suggest that additional public health interventions such as booster vaccine doses, potentially with the more potent vaccine types, may be needed to control the COVID-19 pandemic in Mongolia and worldwide.

## Data Availability

Raw data are available upon request.

## Acknowledgements

The authors gratefully acknowledge the study participants for their contributions to this research. The authors thank the laboratories of Jesse Bloom and Peter Kim for providing the pseudovirus plasmid system, and the laboratories of Dennis Burton and Peter Kim for providing HeLa cells expressing hACE2. We thank Maria Filsinger Interrante and Abigail Powell for guidance with the pseudovirus assay, and Javaria Najeeb and Ana Rita Cardoso for helpful discussions. Funding support: NIH/NIAID HHSN272201700013C (GBS); NIH/NIAID R01AI127877 (SDB), NIH/NIAID R01AI130398 (SDB), NIH 1U54CA260517 (SDB, BAP).

