## Supplemental File for "Direct Comparison of Antibody Responses to Four SARS-CoV-2 Vaccines in Mongolia"

**Supplemental Materials**

**Methods**

*Recruitment and Informed Consent of Research Participants*
Research participants were recruited by public announcement. Informed consent for research participation and collection of blood and nasopharyngeal swab specimens was obtained, with research volunteers signing a consent form approved by the Ethics Review Board at the Ministry of Health of Mongolia. Blood specimens were collected in a five-day period from July 3 to 7, 2021. Of the initial 794 participants enrolled in the study, 196 participants balanced according to age, sex and time post-second vaccine dose were selected for serological analysis, with 47, 50, 45 and 54 recipients of the Pfizer/BioNTech, AstraZeneca, Sputnik V and Sinopharm vaccines, respectively. An additional 99 participants who had a history of SARS-CoV-2 infection after vaccination (1, 21, 4 and 73 recipients of the Pfizer/BioNTech, AstraZeneca, Sputnik V and Sinopharm vaccines, respectively) confirmed by real-time PCR or rapid diagnostic testing (RDT), were also studied with serological assays.

*Electrochemiluminescence (ECL) IgG Binding Assay*
Vaccine recipient plasma samples were heat-inactivated at 56°C for 30 minutes. IgG antibodies targeting the Wuhan-Hu1 SARS-CoV-2 nucleocapsid (N), spike (S) and spike receptor binding domain (RBD) were detected using MSD V-PLEX SARS-CoV-2 Panel 4 (IgG) kits (Meso Scale Discovery) according to the manufacturer protocols. The kits measure samples in a 96-well plate by multiplexed indirect serology using patterned arrays of the target antigens and electrochemiluminescence (ECL) detection. Plasma samples were analyzed in duplicate at a 1:5,000 dilution, detected with anti-human IgG antibodies labeled with an ECL label (SULFO-TAG), and quantified with an MSD MESO QuickPlex SQ 120 instrument. In addition to test samples, each plate contained a blank well, three positive control samples, and a 7-point calibration curve in duplicate generated by serial dilution of a reference standard. The calibration sample data for each antigen was fit to a 4-parameter logistic (4-PL) model using 1/Y2 weighting. Antibody unit concentrations (MSD AU/mL) for samples were calculated by backfitting ECL signals to the models.

*ACE2-Variant RBD Antibody Blocking Assays*
Antibodies blocking the binding of ACE2 protein to SARS-CoV-2 RBD for viral variants Alpha (N501Y), Beta (K417N, E484K, N501Y), Gamma (K417T, E484K, N501Y), Delta (L452R, T478K), Epsilon (L452R), Eta/Iota/Zeta (E484K), Kappa (L452R, E484Q), B.1.526.2 (S477N) and P.3 (E484K, N501Y), and Wuhan-Hu1were detected with MSD VPLEX SARS-CoV-2 Panel 11 (ACE2) kits according to the manufacturer's protocols. Heat inactivated plasma samples from vaccinees were analyzed in duplicate at a dilution of 1:10. Samples were added to wells of 96-well plates presenting arrays of the different RBDs and incubated to allow antibodies in the samples to bind. Human ACE2 protein conjugated with the SULFO-TAG label was then added to the wells. After incubating to let labeled ACE2 bind to ACE2-binding sites that were not blocked by antibodies, the plates were read with a MESO QuickPlex SQ 120 instrument. In addition to the test samples, each plate contained a 7-point calibration curve in duplicate generated by serial dilution of a reference standard and a blank well. Results are reported as

percent inhibition calculated based on the equation ((1 – Average Sample ECL Signal / Average ECL signal of blank well) x 100).

*Production of SARS-CoV-2 Pseudotyped lentivirus*
As previously described by Crawford et al. (Crawford et al., 2020), HEK293T cells were transfected via calcium phosphate with a five-plasmid 3rd generation lentiviral system composed of a lentiviral packaging vector (pHAGE_Luc2_IRES_ZsGreen-W), SARS-CoV-2 Wuhan-Hu1 Spike plasmid (pHDM-IDTSpike_fixK), and 3 Helper plasmids containing Gag-Pol (pHDM-Hgpm2), Tat (pHDM-Tat1b), and Rev (pRC-CMV_Rev1b). Briefly, HEK293T cells were plated in 10 cm tissue culture dishes and grown to 70% confluence in DMEM supplemented with 10% FBS, Penicillin/Streptomycin/Glutamine, and 10 mM HEPES. Plasmids were combined in 500 µL of water in the following quantities: 10 µg lentiviral packaging vector, 3.4 µg Spike plasmid, 2.2 µg each Helper plasmid. 500 µL 2X HEPES-Buffered Saline was added dropwise to the DNA mixture followed by 100 µL 2.5 M Calcium Chloride dropwise while agitating the mixture to avoid clumping the DNA. After a 20 min incubation at room temperature, the transfection reaction was added dropwise to the cells with gentle swirling. After 24 h incubation at 37°C, 5% $CO_2$, media was gently aspirated from cells, spun down (300 x g, 5 min) to remove cellular debris, then passed through a 0.45 µm filter, aliquoted, and stored at -80°C.

*Pseudovirus Neutralization Assay*
HeLa cells expressing human ACE2 were plated on the inner 60 wells of a 96-well white-walled, flat clear bottom plates at a density of 5,000 cells per well (100 uL of a 50,000 cell/mL suspension). 200 µL of PBS was placed in the outer wells to reduce evaporation during incubation periods. After 24 h incubation at 37°C, 5% $CO_2$, media was replaced with 100 µL of pseudovirus/serum mixtures, with virus only wells serving as a positive control and media only wells as a negative control. To prepare pseudovirus/serum mixtures, serum was heat inactivated at 56°C for 30 min, diluted 1:25 in media, then serial diluted by 2-fold. Pseudovirus was diluted 1:2 with media and supplemented with Polybrene at 1:1,000 to improve infection efficiency. Serum dilutions and pseudovirus was then combined 1:1 for a final starting dilution of 1:50. After 48 h incubation, plates were read by replacing 50 µL of pseudovirus/serum mixture with 50 µL Perkin Elmer BriteLite Plus Luciferase reagent. Plates were read by BioTek Synergy 2 plate reader.

*Statistical Analysis for Pseudovirus Neutralization Assays*
Luciferase readout values were normalized by the average positive and negative control values on each sample's respective plate. IC50 values were calculated by 4 point non-linear regression with a constraint of 0% set to the bottom of the fit and 100% for the top of the fit.

*SARS-CoV-2 Variant Genotyping*
Total nucleic acids were extracted from 182 SARS-CoV-2 positive nasopharyngeal swab specimens (300 µL) using the Chemagic Viral DNA/RNA 300 Kit on the Chemagic 360 extraction instrument (both from Perkin-Elmer, Waltham, MA) according to the manufacturer's instructions.

Purified nucleic acid eluates were genotyped using a multiplex, mutation-specific RT-qPCR targeting N501Y, E484K, and L452R, as previously described (PMID: 34037430). A second confirmatory genotyping RT-qPCR assay was then used to identify the Alpha variant in N501Y mutation-positive samples (n=177) from the first multiplex reaction. For this assay we designed primers and a dual-labeled hydrolysis probe targeting spike del69_70 (Table S1). The N501Y mutation was also included as a positive control, as this Alpha variant confirmation assay was run only on samples positive for N501Y in the first reaction. The del69_70 mutation was selected for Alpha variant confirmation given that 99.93% (929,411/930,076) of adequately covered [unidentified nucleotides (N) <5%], full-length SARS-CoV-2 sequences in GISAID (as of June 22, 2021) with N501Y and del69_70 belonged to the Alpha variant B.1.1.7 lineage. Primers and probes for the spike P681H mutation were also included in the reaction, but this target was not utilized in this study.

Primer/probe mix (1 µL, final concentration 360 nM each primer, 80 nM each probe) was combined with a one-step RT-qPCR system (12.5 µL master mix + 0.5 µL Taq polymerase, SuperScript™ III Platinum™ One-Step qRT-PCR Kit, Invitrogen, Carlsbad), nuclease-free water (15 µL), and template (5.0 µL) in a 25 µL reaction. All experiments were conducted on a BioRad CFX96 real-time PCR instrument (BioRad, Hercules, CA, USA). One mutant control (pooled mutant ssDNA) and one wild-type control (whole-genome synthetic RNA from Twist Bioscience, South San Francisco, CA) were included in each RT-qPCR experiment. Cycling conditions were as follows: 52°C for 15:00, 94°C for 2:00, and then 45 cycles of 94°C for 00:15, 59.0°C for 00:40, and 68°C for 00:20. Fluorescence thresholds were manually set at 500 for both N501Y-FAM and del69_70-HEX.

#### Supplemental Figures

#### Figure S1

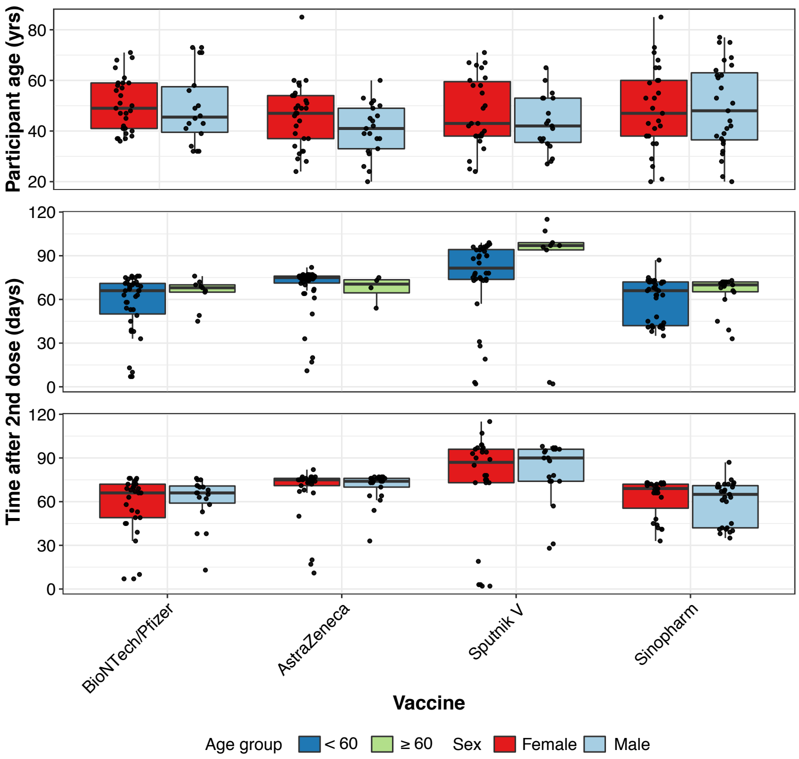


**Vaccine recipient demographics and post-vaccination sampling times.**
(Upper panel) Age of 196 vaccine recipients, stratified by sex.
(Middle panel) Plasma time points sampled after second vaccine dose for participants <60 years old or ≥60 years old.
(Lower panel) Plasma time points sampled for participants stratified by sex.

#### Figure S2

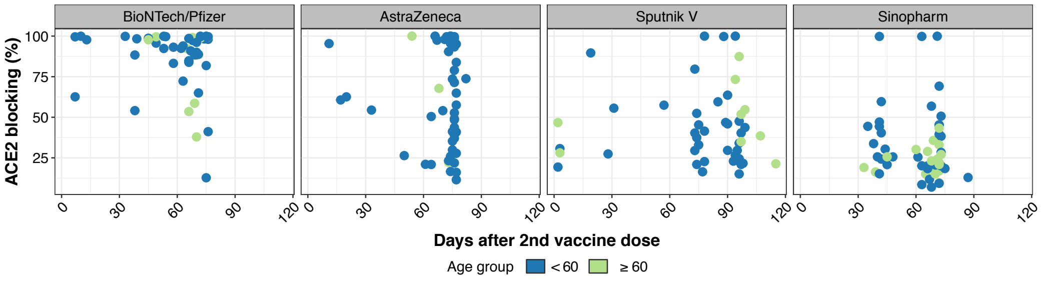


**Vaccine-induced antibody blocking of RBD-ACE2 binding as function of time post-vaccine second dose.** RBD-ACE2 blocking percentages are shown for plasma time points sampled after second vaccine dose for participants stratified by age group (<60 years old or ≥60 years old).

### Figure S3

**A**

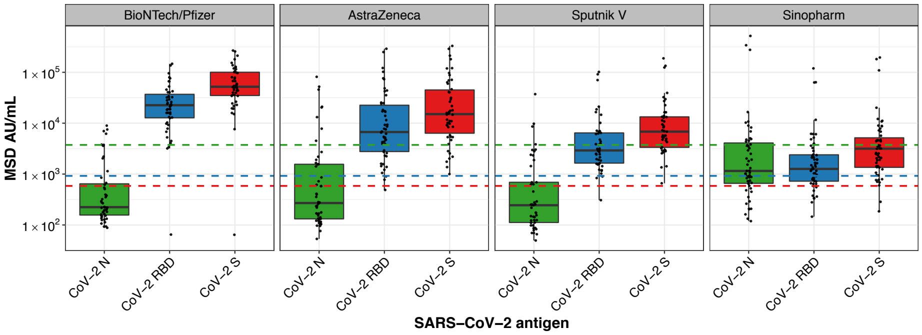


**B**

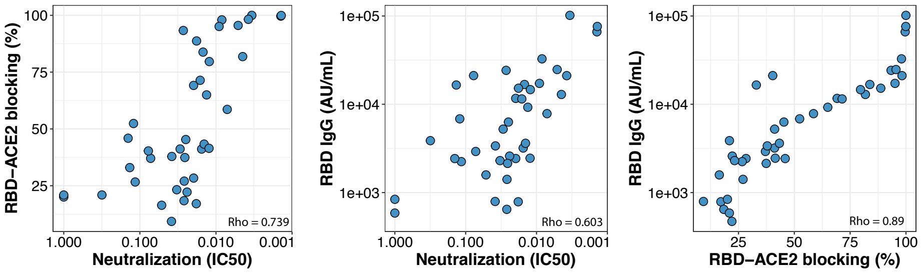


**Concentrations of IgG antibodies specific for SARS-CoV-2 proteins, and correlations with ACE2 blocking and pseudotyped neutralization assays with Wuhan-Hu1 antigens**. (**A**) SARS-CoV-2 nucleocapsid (N), RBD and spike (S)-binding IgG concentrations in vaccine recipient plasmas. Dashed lines show the cut-off values for positive test results for each antigen (green=nucleocapsid, blue=RBD, red=spike). (**B**) Correlations between RBD-ACE2 blocking antibody assay, lentiviral pseudotyped neutralization assay, and anti-RBD IgG results. Spearman's Rho for correlation of RBD-ACE2 blocking and neutralization was 0.739, for RBD IgG and neutralization was 0.603, and for RBD IgG and RBD-ACE2 blocking was 0.89.

#### Figure S4

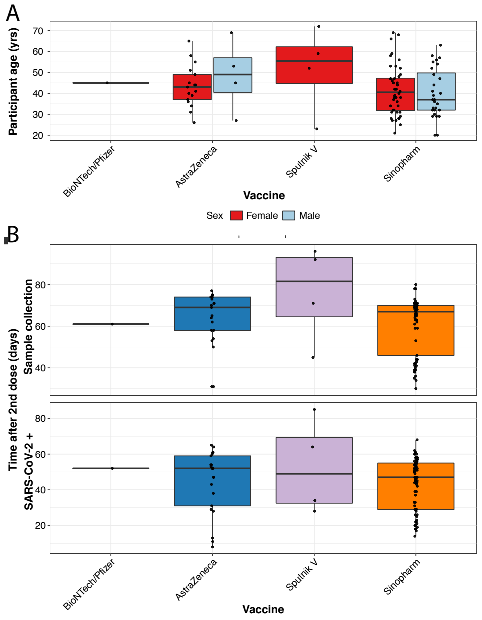


**Demographics as well as post-vaccination sampling and SARS-CoV-2 infection times.**
(A) Age of 99 vaccine recipients with breakthrough infections, stratified by sex.
(B) Plasma time points sampled after second vaccine dose (upper panel) and SARS-CoV-2 infection time points after second vaccine dose (lower panel).

**Supplemental Table 1**. Primer and probe sequences and concentrations for viral variant-specific RT-qPCR assays.

| Oligonucleotide | Sequence (5' to 3') | 5' Mod | 3' Mod | Final reaction concentration |
|---|---|---|---|---|
| N501Y_FWD | GTTTTAATTGTTACTTTCCTTTACAATC | - | - | 360 nM |
| N501Y_REV | CTTTTTAGGTCCACAAACAGT | - | - | 360 nM |
| N501Y_MT_FAM | TTTCCAACCCACTTATGGT | FAM | BHQ-1 | 80 nM |
| del69_70_FWD | CATTAAATGGTAGGACAGGGTTA | - | - | 360 nM |
| del69_70_REV | ACATTCAACTCAGGACTTGTT | - | - | 360 nM |
| del69_70_MT_HEX | TTGGTCCCAGAGATAGCATG | HEX | BHQ-1 | 80 nM |
| P681H_FWD | CAGGTATATGCGCTAGTTATCAG | - | - | 360 nM |
| P681H_REV | CACCAAGTGACATAGTGTAGG | - | - | 360 nM |
| P681H_MT_Cy5 | CAGACTAATTCTCATCGGCG | Cy5 | BHQ-2 | 80 nM |

FAM, 6-Carboxyfluorescein; HEX, Hexachloro-Fluorescein; Cy5, Cyanine-5; BHQ, Black Hole Quencher
